# Age and Sex-Based Differences in Proctological Diseases: A Cross Sectional Analysis of One Thousand and Seven Hundred and Seventy (1770) Anorectoscopy Reports

**DOI:** 10.1101/2025.10.30.25339148

**Authors:** Kaly Keïta, Boua Daoud Camara, Alassane Alfousséni Doumbia, Karim Dao, Cheick Oumar Kamissoko, Drissa Sangaré, Bacary Diarra, Salif Sanafo, Moussa Sangaré, Nouhoum Koné, Sékou Landouré, Mamadou Mallé, Mamadou Cissoko, Sanra Déborah Sanogo, Ganda Soumaré, Moussa Younoussou Dicko, Ibrahima Amadou Dembélé, Djibril Sy, Djénèbou Traoré, Assétou Soukho Kaya, Mamadou Dembélé, Hamar Alassane Traoré

## Abstract

**Background:** The epidemiological characterization of the proctological diseases and the determination of the correlations between the different age group and sex and the proctological diseases may reveal critical information on the distribution of proctological diseases and establish pilot data for age- and sex-specific determinant research, which could lead to adapt preventive and treatment strategies from the decision-makers. We aimed in this study to describe the epidemiological aspects of proctological diseases in digestive endoscopy Unit of internal medicine department at the University Hospital Center of the Point G.

**Methods:** A cross-sectional study with retrospective data collection was conducted between January 01, 2011 to December 31, 2018 from patients who underwent the anorectoscopy in the digestive endoscopy Unit of internal medicine Department at the University Hospital Center of the Point G during the study period.

**Results:** Globally, 1771 reports of anorectoscopy were considered for analysis. The male accounted for 66.2% of cases with a sex-ratio of 1.9. The mean age was 41,17 ± 15,98 years and the extreme ages were 2 years and 75 years. Hemorrhoidal diseases represented 67.3% of cases followed by anitis and anal fissure with 11.7% of cases and 10.3% of cases respectively. The age group under 24 years was 2.03 times and 3.93 times significantly associated with anitis and rectal prolapse (47 cases; OR= 2.03; 95% CI= 1.42 – 2.90; p=0.000) and (8 cases; OR= 3.93; 95% CI= 1.61 – 9.57; p=0.005) respectively. Anal fistula and anal fissure occurred more frequently in age group of 25 – 44 years (36 cases; OR= 1.65; 95% CI= 1.01 – 2.71; p=0.044) and (91 cases; OR= 1.38; 95% CI= 1.02 – 1.88; p=0.038) respectively. The age group of 45 – 59 years was more significantly associated with hemorrhoidal diseases (256 cases; OR= 1.32; 95% CI= 1.02 – 1.70; p=0.032). Anorectal tumor was 1.92 times more likely to be found in of group of 60 – 79 years (23 cases; OR= 1.92; 95% CI= 1.18 – 3.13; p=0.007). Males were 1.65 times more likely than females to develop hemorrhoidal diseases (834 cases vs. 358 cases; OR= 1.65; 95% CI= 1.35 – 2.03; p=0.000). Anal fistula were 2.63 times more likely to be found in males than in females (55 cases vs. 11 cases; OR= 2.63; 95% CI= 1.36 – 5.06; p=0.002).

**Conclusion:** This study shows that the distribution of proctological diseases remains disparate. Anitis and rectal prolapse were significantly associated with the age group under 24 years; the anal fistula and anal fissure with age group of 25 – 44 years; the hemorrhoidal diseases with age group of 45 – 59 years; and the anorectal tumor with age group 60 – 79 years. Males were more likely than females to develop hemorrhoidal diseases and anal fistula.

## BACKGROUND

The distribution of proctological diseases is very disparate according to epidemiological data from proctology literature. Indeed, hemorrhoidal diseases, anal fissures and anorectitis were more frequently found compared to the anal abcess, rectal prolapse, cancer ano-rectal according of these studies [1, 2]. In addition, geographic biases do exist in such distribution of proctological diseases, notably country bias, regional bias and northern-southern bias. Hemorrhoidal diseases were much more frequent in Central African Republic series than Senegalese series [1, 2]. Furthermore, proctological dermatological disorders were higher prevalent in northern areas than southern areas [3 - 6].

Descriptive data revealed that certain proctological diseases occur more frequently in young adult, such as the hemorrhoidal diseases, the anal fissure, and the anal fistula [7 - 9.]. However, a cross sectional analysis of age based variations in all the proctological diseases remains an opened research question in the literature.

The sex-based immunological and hormonological differences leading to the observed differences in disease susceptibity are largely documented [10 - 12]. However, these are less investigated for proctological diseases [6]. Indeed, certain descriptive studies conducted in Mali, in Nigeria, and in France reported the male predominance in hemorrhoidal diseases, female in perianal abscess, and equally distribution in both sexes in anal fissure [7 - 9, 13].

The epidemiological characterization of the proctological diseases and the determination of the correlations between the different age group and sex and the proctological diseases may reveal critical information on the distribution of proctological diseases and establish pilot data for age- and sex-specific determinant research, which could lead to adapt preventive and treatment strategies from the decision-makers.

We aimed to describe the epidemiological aspects of proctological diseases in digestive endoscopy Unit of of internal medicine department at the University Hospital Center of the Point G. The specific objectives included the distribution of patients according to the sociodemographic data. Also, we have determined the panoramic profile of proctological diseases. Finally, we have determined the association between the different age group and sex, and the proctological diseases.

## METHODS

**Research questions:** the study addresses mainly two research questions: i) what are the distribution of the proctological diseases? ii) Are there age- and sex-based differences in proctological diseases?

**Study design:** a cross-sectional study with retrospective data collection was conducted to determine the epidemiological aspects of all proctological diseases and to identify the correlation between the different age groups and sex, and the proctological diseases. This study adhered to the cross-sectional reporting guidelines of Strengthening of Reporting of Observational Studies in Epidemiology (STROBE) [14].

**Study setting:** the study was conducted between January 01, 2011 to December 31, 2018, which is 8 years. It was carried out in the digestive endoscopy Unit of internal medicine Department at the University Hospital Center of the Point G.

**Study population:** we included in this study all the patients who underwent the anorectoscopy in the digestive endoscopy Unit of internal medicine Department at the University Hospital Center of the Point G during the study period and whose the reports of anorectoscopy were exploitable. We did not include the patients who underwent the anorectoscopy outside our department, and the study period and those the reports of anorectoscopy were not exploitable.

**Study materials and procedures**: it was performed in all patients, the collection of the patient’s identity and clinical indications; physical examination of the anal margin in search of internal hemorrhoidal prolapse, associated pathologies (anal fissure, anal fistula), external hemorrhoidal thrombosis; digital rectal examination to assess the anal sphincter tonicity ; to look for localized pain, to visualize a blood-stained finger cot and to eliminate an anorectal tumor mass, anoscopy for visual exploration of the anal canal and internal hemorrhoid diseases; rectoscopy for visual exploration of the rectal mucosa up to 20 centimeters from the anal margin, and finally the anorectoscopy report. In addition, internal hemorrhoidal pathology was classified into 4 grades of increasing severity [15]: Grade 1 visible on proctoscopy as a bulging vascular cushion; Grade 2 at times prolapse during defecation, then spontaneous reduction; Grade 3 prolapse during defecation, no spontaneous reduction, only digitally reducible; Grade 4 nonreducible hemorrhoidal prolapse. Complications such as internal thrombosis, cryptitis, papillitis, polypoid formations were investigated.

**Variables:** the independent study variables were the socio-demographic information which included sex, age, ethnic group, profession, and residence. The proctological diseases are the dependent (outcome) variables, for which diagnoses were established using an anorectoscopy examination.

## Data sources/ measurement

**Data collection tool:** a pre-established survey form was designed and used to collect on the sociodemographic and the results of anorectoscopy.

**Data collection:** sociodemographic variables such as sex, age, profession, residence and the results of anorectoscopy were collected from the registry of anorectoscopy in which there are the reports of anorectoscopy.

**Difficulties and biases:** we encountered, during this study, some problems related to the lack of information in the reports of anorectoscopy. Both the lower digestive endoscopist and the prescriber did not inform correctly sometime the reports of anorectoscopy and the clinical indications, respectively.

**Sampling and sample size:** this was an exhaustive sampling of all cases of the proctological diseases mentioned in the registry of anorectoscopy during the study period. The sample size was not calculated.

**Statistical methods:** data entry and analysis were done using SPSS version 22 software. Data cleaning was done by checking and correcting for duplicates and completing missing data, and correcting outliers. We conducted statistical analyses using Epi Info version 7.2 and SPSS version 22 software. We used Microsoft Excel to generate bar graphs. Quantitative data were presented as mean and standard deviation such as the age. Qualitative data were presented as numbers and percentages. In the bivariate analysis, we calculated odds ratios (OR), 95% confidence intervals (C.I), and p-values. The outcome variables of interest for bivariate analysis were proctological diseases. The categorical independent variables of interest were age group and sex. The Chi-square (uncorrected and corrected) and Fisher‘s exact tests were used to assess the statistical significance and strength of the associations between the categorical independent variables and the outcome variables. A two-tailed p-value <0.05 were retained and considered as statistically significant.

**Ethical consideration:** according to Helsinki guideline, research involving human subject should be conducted ethically, with the well-being of the subject taking priority over scientific or societal interests. Our research study aiming to provide a cross-sectional analysis of one thousand and seven hundred and seventy (1770) anorectoscopy reports in order to explore the age and sex-based differences in proctological diseases involved human subjects. However, we used secondary data notably data extraction on the registry of anorectoscopy (sociodemographic data and anorectoscopy findings), but not the biological specimens. In addition, the study was retrospective and all data was extracted anonymously from the registry of anaorectoscopy. Therefore, patients’ informed consent was not required. Given the nature of this study, formal ethical approval from an ethics committee was not sought. However, formal permission to conduct this study was obtained from the General Director of University Hospital Center of the Point G. The registry of anorectoscopy was returned in the archive room immediately after exploitation.

## RESULTS

### Characteristics of the study participants

During the study period from January 01, 2011 to December 31, 2018, i.e. 8 years, 1788 anorectoscopies were performed in the digestive endoscopy Unit of internal medicine Department at the University Hospital Center of the Point G. Eighteen reports of anorectoscopy were not exploitable. One thousand and seven hundred and seventy reports of anorectoscopy were considered for analysis. The distribution of patients according to the year is mentioned in figure 1. In 2014, the annual realization rate of anorectoscopy was 16, 8% of cases. The mean annual realization rate of anorectscopy was 221.25 cases. Reports of anorectoscopy were normal in 9.8% cases. The distribution of patients by sociodemographic data is summarized Table 1 and table 1 (continued). The male accounted for 66.2% of cases with a sex-ratio of 1.9. The age group of 25 – 44 years represented 42,5% of cases. The mean age was 41,17 ± 15,98 years and the extreme ages were 2 years and 75 years. The Bamanan ethnic group represented 18,3% of cases followed by Peulh and Malinké ethnic groups with 13.8% of cases and 6.2% of cases respectively. The civil servant accounted for 17.3% of the study population followed by housewife and pupil/student with 15.6% of cases and 10.6% of cases respectively. The majority of patients came from Bamako in 44.9% of cases.

**Table 1.** Distribution of patients by the sociodemographic data.

| **Table 1: Distribution of patients by the sociodemographic data** |  |  |
| --- | --- | --- |
| **Sociodemographic data** | **Number of cases** | **Percentage** |
| **Sex** |  |  |
| Male | 1172 | 66.2 |
| Female | 598 | 33.8 |
| **Age group** |  |  |
| 15 - 24 years | 245 | 13.8 |
| 25 - 44 years | 753 | 42.5 |
| 45 - 59 years | 368 | 20.8 |
| 60 - 79 years | 259 | 14.6 |
| > 80 years | 11 | 0.6 |
| No information | 134 | 7.6 |
| **Ethnic group** |  |  |
| Bamanan | 324 | 18.3 |
| Malinké | 110 | 6.2 |
| Peulh | 245 | 13.8 |
| Bobo | 17 | 1.0 |
| Dogon | 54 | 3.1 |
| Senofé | 23 | 1.3 |
| Sonrai | 60 | 3.4 |
| Soninké | 92 | 5.2 |
| No information | 772 | 43.6 |
| Others | 73 | 4.1 |

| **Table 1 (continued):** Distribution of patients by the sociodemographic data |  |  |
| --- | --- | --- |
| **Sociodemographic data** | **Number of cases** | **Percentage** |
| **Profession** |  |  |
| Housewife | 277 | 15.6 |
| Pupil/Student | 187 | 10.6 |
| Trader | 152 | 8.6 |
| Farmer | 100 | 5.6 |
| Worker | 73 | 4.1 |
| Civil servant | 307 | 17.3 |
| Military | 172 | 9.7 |
| Not employed | 3 | 0.2 |
| No information | 316 | 17.9 |
| Others | 183 | 10.3 |
| **Residence** |  |  |
| Bamako | 794 | 44.9 |
| Outside of the Bamako | 772 | 43.7 |
| Outside of the Mali | 74 | 4.2 |
| No information | 130 | 7.3 |

**Figure 1:**
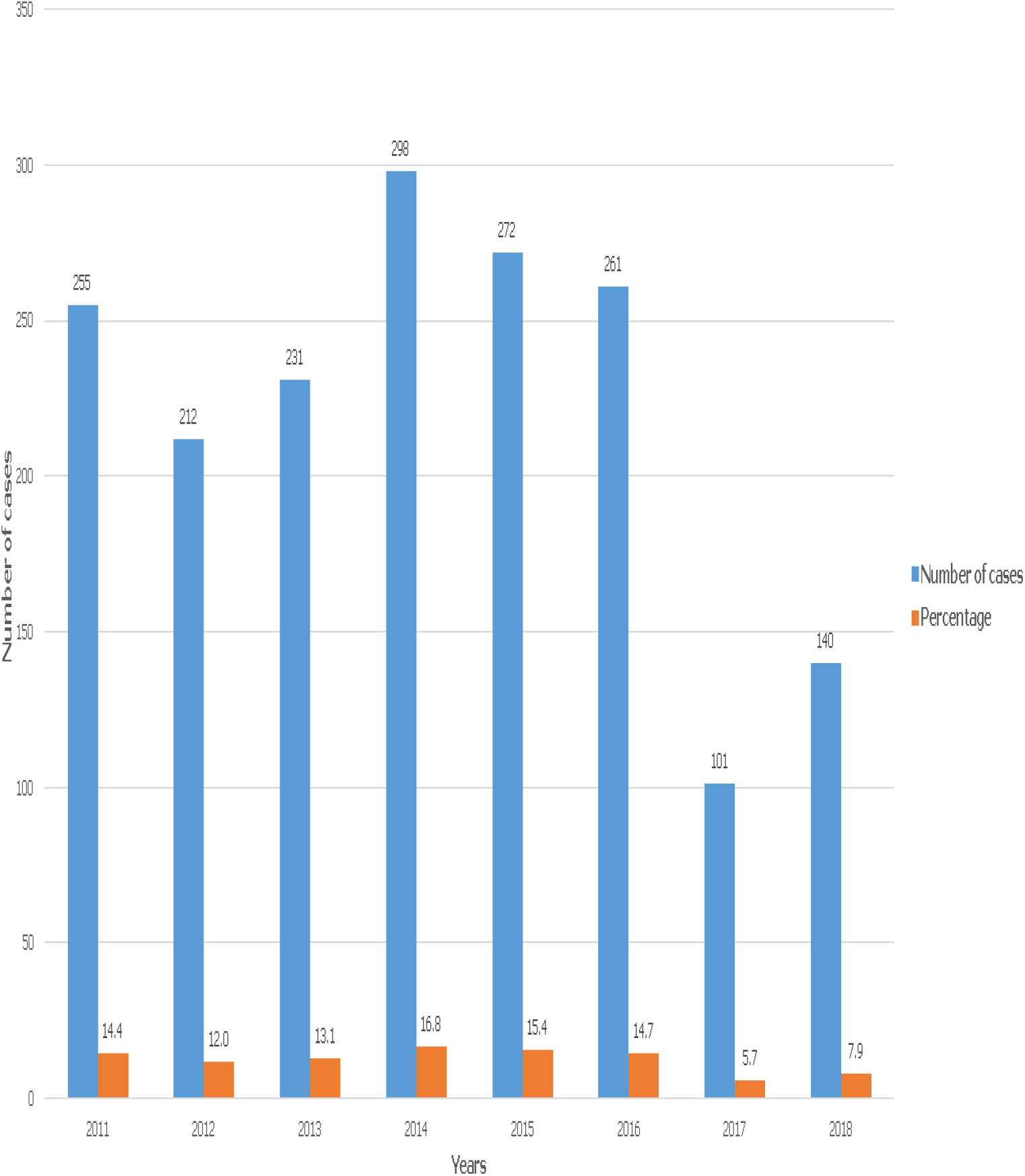
Distribution of patients according to the years of anorectoscopy realization

### Descriptive analysis

The hemorrhoidal syndrome motivated the realization of anorectoscopy with 26.0% of cases followed by rectorragy (16.3% of cases) and anal pain (14.5% of cases), as mentioned in table 2. The table 3 shows the distribution of patients by proctological diseases. Hemorrhoidal diseases represented 67.3% of cases followed by anitis and anal fissure with 11.7% of cases and 10.3% of cases respectively. The characteristics of hemorrhoidal diseases are illustrated in table 4. Out of the 1192 cases of hemorrhoidal diseases, the stage 2 were found in 50.1% of cases, internal hemorrhoidal disease in 82.4% of cases, the thrombosed internal hemorrhoidal disease in 8.5% of cases and the thrombosed external hemorrhoidal disease in 3.7% of cases.

**Table 2.** Distribution of patients according to the clinical indication of anorectoscopy.

|  |  |  |
| --- | --- | --- |
| **Clinical indications** | **Number of cases (N= 1770)** | **Percentage** |
| Hemorrhoidal syndrome<sup>a</sup> | 461 | 26.0 |
| Rectorragy | 288 | 16.3 |
| Anorectal clinical mass syndrome<sup>b</sup> | 79 | 4.5 |
| Extension work up of a primary cancer | 60 | 3.4 |
| Constipation | 41 | 2.3 |
| Anal pain | 256 | 14.5 |
| Anal pruritus | 25 | 1.4 |
| Anal fissure | 48 | 2.7 |
| No information | 72 | 4.1 |
| Anal fistula | 52 | 2.9 |
| Anoperineal tumefaction | 57 | 3.2 |
| Others | 331 | 18.7 |
| Total | 1770 | 100.0 |

<sup>a</sup>Hemorrhoidal syndrome: all the hemorrhoid related symptoms that motivated the realization of anorectoscopy for which prescriber noted suspicion of hemorrhoid as clinical information.
<sup>b</sup>Anorectal clinical mass syndrome: all the anorectal tumor related symptoms for which prescriber noted suspicion of anorectal tumor.

**Table 3.** Distribution of patients according to the proctological diseases.

|  |  |  |
| --- | --- | --- |
| **Proctological diseases** | **Number of cases (N= 1770)** | **Percentage** |
| Hemorrhoidal diseases | 1192 | 67.3 |
| Anitis | 207 | 11.7 |
| Anal fissure | 183 | 10.3 |
| Anorectal tumor | 96 | 5.4 |
| Intergluteal mycosis | 74 | 4.2 |
| Anal fistula | 66 | 3.7 |
| Rectitis | 44 | 2.5 |
| Anorectitis | 42 | 2.4 |
| Anoperineal abcess | 24 | 1.4 |
| Anal ulcer/ulceration | 16 | 0.9 |
| Rectal prolapse | 21 | 1.2 |
| Other proctological abnormalities | 622 | 35.1 |

Other proctological diseases: hemorrhoidal marisk (579 cases/1770), anal eczema (16 cases/1770), Melena (5 cases/1770), extrensic compression of the rectal (14 cases/1770), Fecaloma (13 cases/1770), Anal beance (2 cases/1770), anal sphincter hypotony (2 cases/1770), post-surgery narrowed anal canal (2 cases/1770), polyps (18 cases/1770), prostatic hypertrophy (17 cases/1770).

**Table 4.** Distribution of patients according to the hemorrhoidal disease characteristics.

|  |  |  |
| --- | --- | --- |
| **Hemorrhoidal disease characteristics** | **Number of cases (n= 1192)** | **Percentage** |
| **Stage of hemorrhoidal diseases** |  |  |
| Stage I | 390 | 32.7 |
| Stage II | 597 | 50.1 |
| Stage III | 77 | 6.5 |
| Stage IV | 6 | 0.5 |
| No information | 56 | 4.7 |
| **Type of hemorrhoidal diseases** |  |  |
| Internal | 982 | 82.4 |
| External | 66 | 5.5 |
| Internal and external | 144 | 12.1 |
| **Thrombosed internal hemorrhoidal diseases** |  |  |
| Yes | 101 | 8.5 |
| No | 1091 | 91.5 |
| **Thrombosed external hemorrhoidal diseases** |  |  |
| Yes | 44 | 3.7 |
| No | 1148 | 96.3 |

### Age-based differences in proctological diseases

The distribution of patients according the age groups and the proctological diseases is mentionned in table 5 and table 5 (continued). The age group under 24 years was 2.03 times and 3.93 times significantly associated with anitis and rectal prolapse (47 cases; OR= 2.03; 95% CI= 1.42 – 2.90; p=0.000) and (8 cases; OR= 3.93; 95% CI= 1.61 – 9.57; p=0.005) respectively. Anal fistula and anal fissure occurred more frequently in age group of 25 – 44 years (36 cases; OR= 1.65; 95% CI= 1.01 – 2.71; p=0.044) and (91 cases; OR= 1.38; 95% CI= 1.02 – 1.88; p=0.038) respectively. The age group of 45 – 59 years was more significantly associated with hemorrhoidal diseases (256 cases; OR= 1.32; 95% CI= 1.02 – 1.70; p=0.032). Anorectal tumor was 1.92 times more likely to be found in of group of 60 – 79 years (23 cases; OR= 1.92; 95% CI= 1.18 – 3.13; p=0.007).

**Table 5.**
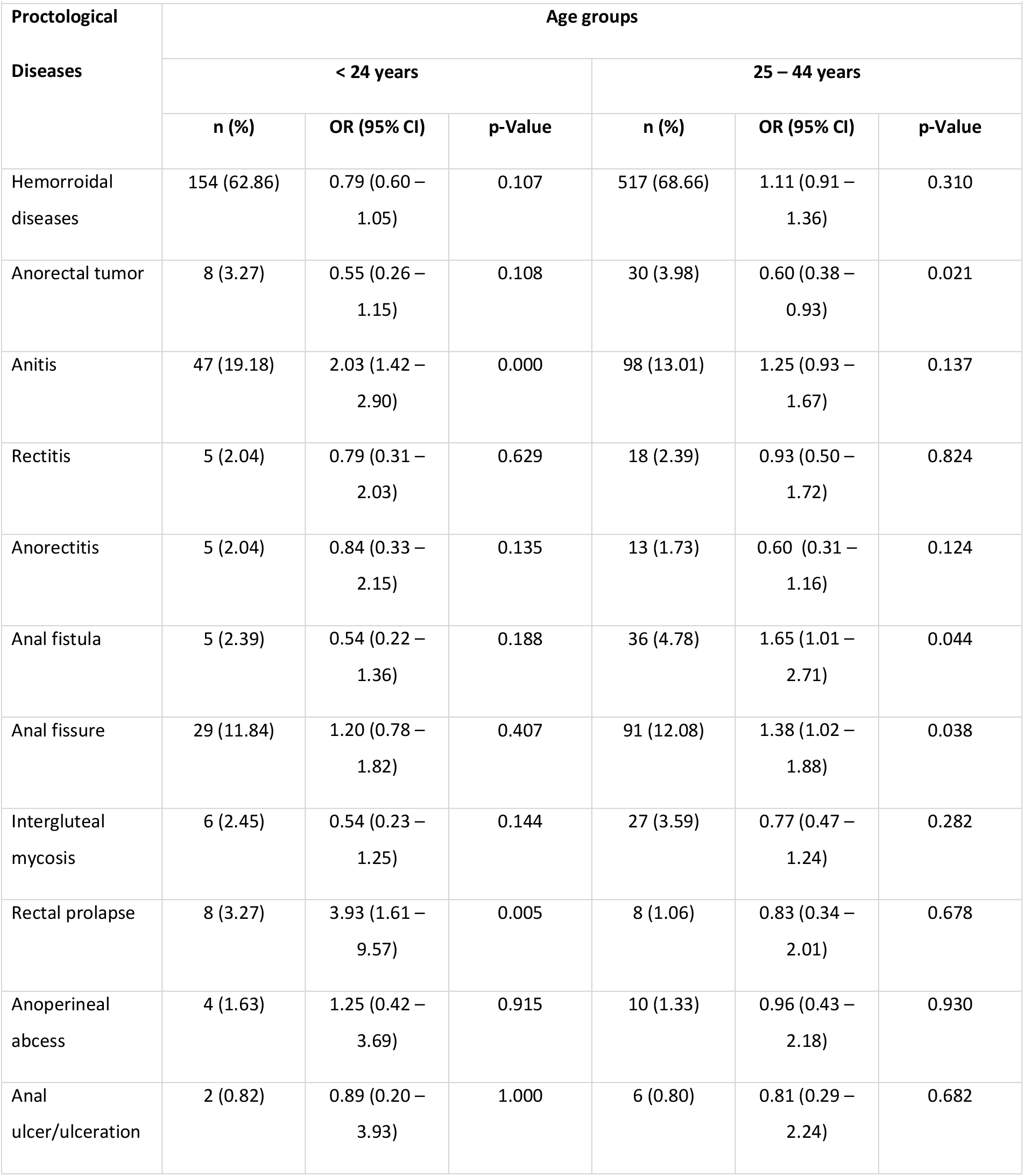
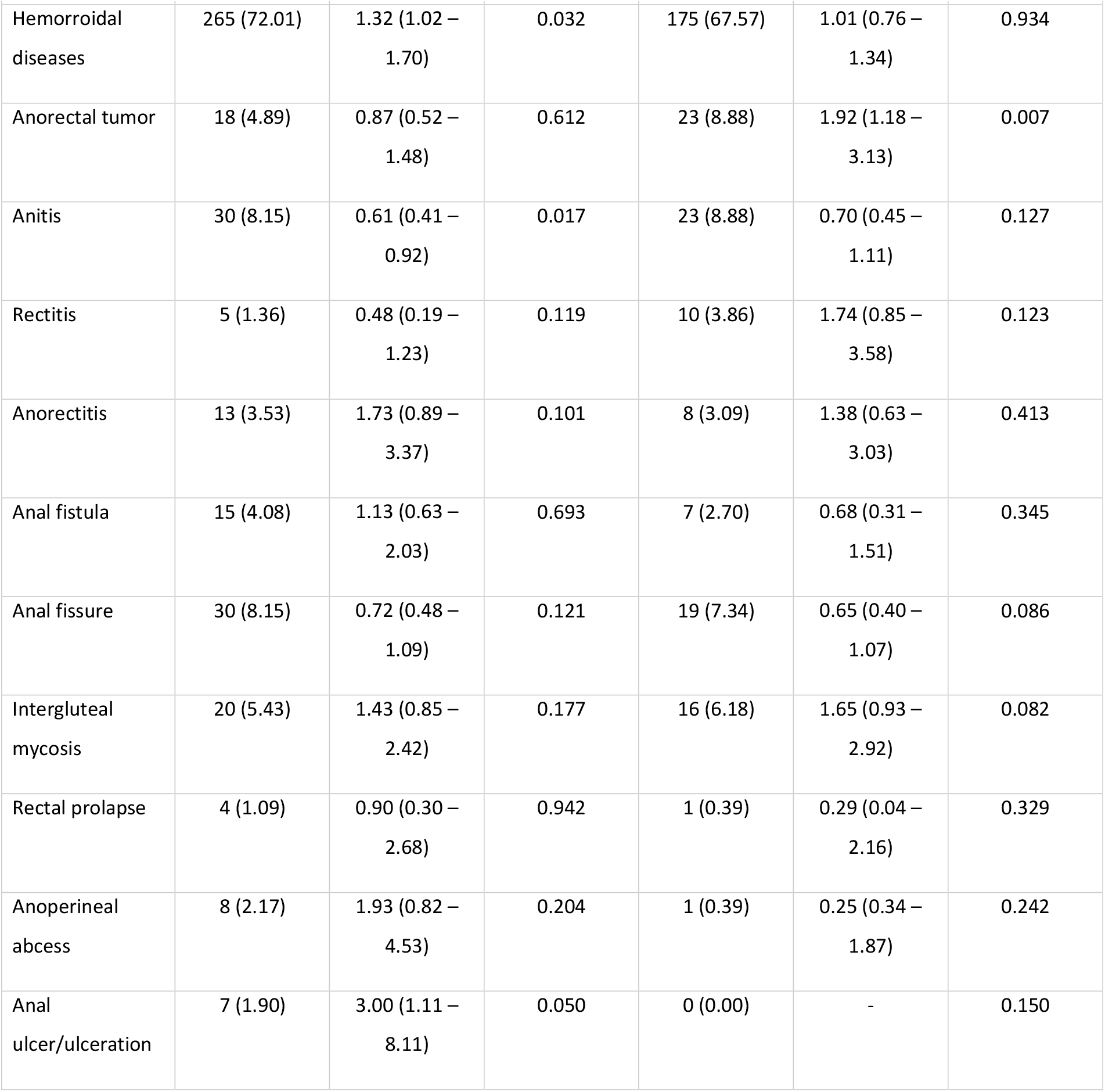
Distribution of patients according to the age group and the proctological diseases.

| Proctological Diseases | Age groups |  |  |  |  |  |
| --- | --- | --- | --- | --- | --- | --- |
|  | < 24 years |  |  | 25 – 44 years |  |  |
|  | n (%) | OR (95% CI) | p-Value | n (%) | OR (95% CI) | p-Value |
| Hemorrhoidal diseases | 154 (62.86) | 0.79 (0.60 – 1.05) | 0.107 | 517 (68.66) | 1.11 (0.91 – 1.36) | 0.310 |
| Anorectal tumor | 8 (3.27) | 0.55 (0.26 – 1.15) | 0.108 | 30 (3.98) | 0.60 (0.38 – 0.93) | 0.021 |
| Anitis | 47 (19.18) | 2.03 (1.42 – 2.90) | 0.000 | 98 (13.01) | 1.25 (0.93 – 1.67) | 0.137 |
| Rectitis | 5 (2.04) | 0.79 (0.31 – 2.03) | 0.629 | 18 (2.39) | 0.93 (0.50 – 1.72) | 0.824 |
| Anorectitis | 5 (2.04) | 0.84 (0.33 – 2.15) | 0.135 | 13 (1.73) | 0.60 (0.31 – 1.16) | 0.124 |
| Anal fistula | 5 (2.39) | 0.54 (0.22 – 1.36) | 0.188 | 36 (4.78) | 1.65 (1.01 – 2.71) | 0.044 |
| Anal fissure | 29 (11.84) | 1.20 (0.78 – 1.82) | 0.407 | 91 (12.08) | 1.38 (1.02 – 1.88) | 0.038 |
| Intergluteal mycosis | 6 (2.45) | 0.54 (0.23 – 1.25) | 0.144 | 27 (3.59) | 0.77 (0.47 – 1.24) | 0.282 |
| Rectal prolapse | 8 (3.27) | 3.93 (1.61 – 9.57) | 0.005 | 8 (1.06) | 0.83 (0.34 – 2.01) | 0.678 |
| Anoperineal abcess | 4 (1.63) | 1.25 (0.42 – 3.69) | 0.915 | 10 (1.33) | 0.96 (0.43 – 2.18) | 0.930 |
| Anal ulcer/ulceration | 2 (0.82) | 0.89 (0.20 – 3.93) | 1.000 | 6 (0.80) | 0.81 (0.29 – 2.24) | 0.682 |
|  | 45 - 59 years, |  |  | 60 – 79 years |  |  |
|  | n (%) | OR (95% CI) | p-Value | n (%) | OR (95% CI) | p-Value |
| Hemorroidal diseases | 265 (72.01) | 1.32 (1.02 – 1.70) | 0.032 | 175 (67.57) | 1.01 (0.76 – 1.34) | 0.934 |
| Anorectal tumor | 18 (4.89) | 0.87 (0.52 – 1.48) | 0.612 | 23 (8.88) | 1.92 (1.18 – 3.13) | 0.007 |
| Anitis | 30 (8.15) | 0.61 (0.41 – 0.92) | 0.017 | 23 (8.88) | 0.70 (0.45 – 1.11) | 0.127 |
| Rectitis | 5 (1.36) | 0.48 (0.19 – 1.23) | 0.119 | 10 (3.86) | 1.74 (0.85 – 3.58) | 0.123 |
| Anorectitis | 13 (3.53) | 1.73 (0.89 – 3.37) | 0.101 | 8 (3.09) | 1.38 (0.63 – 3.03) | 0.413 |
| Anal fistula | 15 (4.08) | 1.13 (0.63 – 2.03) | 0.693 | 7 (2.70) | 0.68 (0.31 – 1.51) | 0.345 |
| Anal fissure | 30 (8.15) | 0.72 (0.48 – 1.09) | 0.121 | 19 (7.34) | 0.65 (0.40 – 1.07) | 0.086 |
| Intergluteal mycosis | 20 (5.43) | 1.43 (0.85 – 2.42) | 0.177 | 16 (6.18) | 1.65 (0.93 – 2.92) | 0.082 |
| Rectal prolapse | 4 (1.09) | 0.90 (0.30 – 2.68) | 0.942 | 1 (0.39) | 0.29 (0.04 – 2.16) | 0.329 |
| Anoperineal abcess | 8 (2.17) | 1.93 (0.82 – 4.53) | 0.204 | 1 (0.39) | 0.25 (0.34 – 1.87) | 0.242 |
| Anal ulcer/ulceration | 7 (1.90) | 3.00 (1.11 – 8.11) | 0.050 | 0 (0.00) | - | 0.150 |

### Sex-based differences in proctological diseases

Table 6 shows the distribution of patients by the sex and proctological diseases. Males were 1.65 times more likely than females to develop hemorrhoidal diseases (834 cases vs. 358 cases; OR= 1.65; 95% CI= 1.35 – 2.03; p=0.000). Anal fistula were 2.63 times more likely to be found in males than in females (55 cases vs. 11 cases; OR= 2.63; 95% CI= 1.36 – 5.06; p=0.002).

**Table 6.** Distribution of patient according to the sex and the proctological diseases.

| Proctological Diseases | Total, N (%) | Sex |  | OR (95% CI) | p-Value |
| --- | --- | --- | --- | --- | --- |
|  |  | Male, n (%) | Female, n (%) |  |  |
| Hemorrhoidal diseases | 1192 (67.34) | 834 (71.16) | 358 (59.87) | 1.65 (1.35 – 2.03) | 0.000 |
| Anorectal tumor | 96 (5.42) | 61 (5.20) | 35 (5.85) | 0.88 (0.58 – 1.35) | 0.569 |
| Anitis | 207 (11.69) | 144 (12.29) | 63 (10.54) | 1.19 (0.87 – 1.62) | 0.278 |
| Rectitis | 44 (2.49) | 32 (2.73) | 12 (2.01) | 1.37 (0.70 – 2.68) | 0.355 |
| Anorectitis | 42 (2.37) | 21 (1.79) | 21 (3.51) | 0.50 (0.27 – 0.93) | 0.025 |
| Anal fistula | 66 (3.73) | 55 (4.69) | 11 (1.84) | 2.63 (1.36 – 5.06) | 0.002 |
| Anal fissure | 183 (10.34) | 129 (11.01) | 54 (9.03) | 1.25 (0.89 – 1.74) | 0.196 |
| Intergluteal mycosis | 74 (4.18) | 43 (3.67) | 31 (5.18) | 0.70 (0.43 – 1.12) | 0.132 |
| Rectal prolapse | 21 (1.19) | 14 (1.19) | 7 (1.17) | 1.02 (0.41 – 2.54) | 0.965 |
| Anoperineal abcess | 24 (1.36) | 16 (1.37) | 8 (1.34) | 1.02 (0.43 – 2.39) | 0.962 |
| Anal ulcer/ulceration | 16 (0.90) | 13 (1.11) | 3 (0.50) | 2.22 (0.63 – 7.84) | 0.201 |

## DISCUSSION

### Main findings

During this 8-year period study on 1788 anorectoscopies performed in the digestive endoscopy Unit of internal medicine Department at the University Hospital Center of the Point G of which eighteen reports of anorectoscopy were not exploitable and whose one thousand and seven hundred and seventy reports of anorectoscopy were finally considered for analysis have confirmed that the distribution of proctological diseases remain disparate as previous panoramic studies [1, 2, 4 – 6, 16]. In addition, this study also demonstrates that there are age and sex based differences in some proctological diseases.

### Characteristics of the study participants

Of 1788 anorectoscopies performed in the digestive endoscopy Unit of internal medicine Department at the University Hospital Center of the Point G, the higher annual realization rate of anorectoscopy was observed in 2024. The mean annual realization rate of anorectoscopy was 221.25 cases. The large variations observed between these annual realization rates of anorectoscopy could be explain by the fact that there were frequently problems with anorectoscopy tools and the untimely strike by hospital personnel.

Most of epidemiological studies on proctological diseases shows the male predominance [1, 2, 4, 17], in accordance with our study findings. However, Abramowitz et al. studied the prevalence of proctological symptoms amongst patients who see general practitioners in France, in which authors reported the female predominance [3]. This discrepancy may could be explained by the fact that ours studies were hospital-based study compared to study conducted by Abramowitz et al. in France, which was population-based study.

Several data from published works indicate that the proctological diseases occurred more frequently in young adults [1, 2, 18], as were found in our study. In contrast, Abramowitz et al. noted a mean age of 54 years, which were above to ours [3]. The high average life expectancy in western setting, where Abramowitz et al. had conducted their study, which is largely above to ours. This may explain these differences between the studies.

In our study, the bamanan ethnic group was more represented followed by peulh and malinké ethnic group, this finding parallels with data from the Demographic and Health Survey of Mali (EDSM VI) [19].

The civil servant followed by housewife and pupil/student were more represented in our study population, which was similar to that of another Malian study [20], but not consistent with this previous study [21]. This difference could be due to the methodological approach notably study setting and studied population, which are different.

The majority of patients came from Bamako, in agreement with the study conducted in Mali by Diarra et al. [7]. Both studies were conducted in Bamako, which may explain this result.

### Descriptive analysis

Several studies revealed that clinical indications motivating the realization of anorectoscopy were mainly the anal pain, rectorrhagy, and suspicion of hemorrhoidal diseases, this is in accordance with our study finding [1, 2, 4, 18].

Most of epidemiological studies on proctological diseases that the hemorrhoidal diseases were as far as prevalent than other proctological diseases, as were found in our study [1 – 4, 16].

In our study, half of patients were diagnosed at hemorrhoidal diseases stage 2, most were internal hemorrhoidal diseases and less one to four presented thrombosed internal or external hemorrhoidal diseases, this result parallels with previous study conducted in Mali by Diarra et al. [7].

### Age-based differences in proctological diseases

The results from our study show that anitis and rectal prolapse were signicantly associated with the age group under 24 years; the anal fistula and anal fissure with age group of 25 – 44 years; the hemorrhoidal diseases with age group of 45 – 59 years; and the anorectal tumor with age group 60 – 79 years, not consistent with study conducted in Tchad, which was a rare study that studying univariately correlation between age and proctological diseases. In this study, there were only significant association between anal fissure and age group under 44 years, and colorectal cancer and age group over 44 years [6]. This discrepancy may be explained by methodological approach notably the study population is stratified in two age group in this study, but these were five age groups in our study.

### Sex-based differences in proctological diseases

Mahamat et al. reported that males were more likely to develop hemorrhoidal diseases and colorectal cancer than female [6]. In contrast, in our study, Males were more likely than females to develop hemorrhoidal diseases and anal fistula were more likely to be found in males than in females but there was no statistically significant association between colorectal cancer and sex. The small sample size in the Tchadian study could explain this difference.

### Limitations

Our study has limitations. Firstly, the anorectoscopy performed outside of our study setting, which may have led to under-ascertainment on the latter. Secondly, the lack of some informations on reports of anorectoscopy, which may cause confounding biases. Finally, the study was a single-center study, which may lead a generalization bias.

### Strengths

This study contains several strengths. First, it is a long-term observational study with all reports of anorectoscopy. Second, consent procedures were not required for enrollment which may increasing our sample size. Third, our study demonstrates the scope of the panoramic profile of proctological diseases in internal medicine. Fourth, our data provide significant correlations between certain age groups and sex and the proctological diseases. Finally, it also provides sufficient grounds to more explore the correlation between the age group- and sex-specific determinants and the proctological diseases.

## CONCLUSION

This study shows that the distribution of proctological diseases remains disparate. Proctological diseases are as far as dominated by hemorrhoidal diseases notably internal hemorrhoidal diseases and stage 2 hemorrhoidal diseases at moment of diagnosis. Hemorrhoidal syndrome frequently motivates the realization of anorectoscopy. Anitis and rectal prolapse were signicantly associated with the age group under 24 years; the anal fistula and anal fissure with age group of 25 – 44 years; the hemorrhoidal diseases with age group of 45 – 59 years; and the anorectal tumor with age group 60 – 79 years. Males were more likely than females to develop hemorrhoidal diseases and anal fistula.

## Data Availability

All data produced in the present work are contained in the manuscript.

## ACKNOWLEDGMENTS

We express our sincere gratitude to the head nurse of the Digestive Endoscopy Unit of our department of internal medicine responsible of registry of anorectoscopy and all the healthcare providers who participated in this study.

## AUTHORS’ CONTRIBUTIONS

**Conception:** Kaly Keïta and Hamar Alassane Traoré. **Design of the study:** Kaly Keïta, Boua Daoud Camara, Alassane Alfousséni Doumbia, and Mamadou Dembélé. **Data collection:** Karim Dao, Cheick Oumar Kamissoko, Drissa Sangaré, Bacary Diarra, Salif Sanafo, Moussa Sangaré, Nouhoum Koné, Sékou Landouré, Mamadou Mallé and Kaly Keïta. **Data analysis and interpretation:** Mamadou Cissoko, Ibrahima Amadou Dembélé, Djibril Sy, Djénèbou Traoré, and Kaly Keïta. **Supervision:** Assétou Soukho Kaya. **Writing original draft:** Kaly Keïta, Boua Daoud Camara, Alassane Alfousséni Doumbia. **Writing review and editing:** Kaly Keïta, Boua Daoud Camara, Alassane Alfousséni Doumbia, Sanra Déborah Sanogo, Ganda Soumaré, Moussa Younoussou Dicko, Ibrahima Amadou Dembélé, Djibril Sy, Djénèbou Traoré, Assétou Soukho Kaya, Mamadou Dembélé and Hamar Alassane Traoré. **Guarantor of the study:** Hamar Alassane Traoré. All the authors have read and agreed to the final manuscript.

